# Age at Menopause and Dementia Risk: Blood-Based Biomarker Evidence from a US National Study

**DOI:** 10.64898/2026.11.12.26367954

**Authors:** Sirena Gutierrez, Michelle R. Caunca, Muqi Guo, Eleanor Hayes-Larson, Sadaf A. Milani, Paola Gilsanz, Jacqueline M. Torres, Kaitlin B. Casaletto

## Abstract

**Importance:** Earlier age at menopause associates with increased later life dementia risk, but the biological links that underlie this relationship remain poorly understood in population-based samples.

**Objective:** To evaluate the relationship between history of menopausal timing and AD/ADRD-related blood-based biomarkers in postmenopausal women.

**Design:** Observational study using the U.S. Health and Retirement Study. AD/ADRD-related biomarkers were measured from venous blood draws conducted in 2016 and released in 2025. We estimated linear and nonlinear associations between spontaneous menopause timing and biomarkers, accounting for sociodemographic and health covariates. We evaluated effect modification by APOE 4 carrier status. Secondary analyses evaluated associations with hysterectomy history.

**Setting:** Population-based.

**Participants:** Community-dwelling post-menopausal women aged 65 years or older.

**Exposure:** Primary: Self-reported spontaneous age of menopause among women with no history of hysterectomy. Secondary: Self-reported history of hysterectomy (oophorectomy history was not assessed).

**Main Outcomes and Measures:** Neurofilament light chain (NfL) and glial fibrillary acidic protein (GFAP) measured in plasma, and phosphorylated tau (p-tau181) and growth differentiation factor 15 (GDF-15), measured in serum, by Simoa or ELLA.

**Results:** Among 1449 women ≥ 65 years, 38.4% self-reported a hysterectomy. Among 893 postmenopausal women without a history of hysterectomy (mean [SD] age at biomarker collection, 75.2 [7.2] years; mean age at menopause, 50.2 [5.2] years), menopausal timing was nonlinearly associated with NfL. Between menopausal ages 45–50, menopause timing was associated with higher NfL (0.010 SD units/year, [95% CI: −0.007, 0.027]), while from menopausal ages 50–55, the association reversed (*b*= −0.031, [95% CI: −0.051, −0.011]). Associations with GFAP or pTau-181 were not significant, but were directionally similar to those observed for NfL. Later menopausal timing linearly associated with lower GDF-15 (*b* = −0.015, [95% CI: −0.027, −0.004]). Self-reported hysterectomy was associated with higher GDF-15 (*b*= 0.118, [95% CI: 0.015, 0.220]), but not other biomarkers. Associations did not significantly differ by APOE ε4 status.

**Conclusions and Relevance:** In a nationally representative study, earlier age at menopause in midlife was associated with later life AD/ADRDs biomarkers reflecting neuroaxonal degeneration and inflammation, underscoring the importance of understanding reproductive aging in dementia risk.

**KEY POINTS:** *Question:* What is the association between menopausal timing and blood-based Alzheimer’s disease and related dementias (AD/ADRD) biomarkers among postmenopausal women 65 years and older in the United States?

*Findings:* Menopausal timing is associated with higher NfL in women who experienced menopause before age 50, but lower NfL in women with menopausal ages after 50. Earlier menopausal timing is linearly associated with higher GDF-15. Secondary analyses showed associations between hysterectomy history and higher GDF-15.

*Meaning:* Younger menopausal timing associates with higher AD/ADRD-related biomarkers measured decades later in a population-based cohort, underscoring a link between reproductive aging and future neurodegenerative risk.

## INTRODUCTION

Women comprise approximately two-thirds of individuals living with Alzheimer’s disease (AD) worldwide.^1,2^ Sex-specific biological aging processes, such as menopause, may contribute to this elevated risk. Menopause (average age 51-52 in the U.S.) is characterized by steep declines in endogenous estradiol and progesterone production as well as a rise in gonadotropin hormones (e.g., follicle-stimulating hormone), the latter of which may be neurotoxic and linked to greater AD pathology.^3,4^ Earlier menopause is associated with a host of adverse later-life health outcomes,^5,6^ including higher dementia risk.^7–13^ A recent meta-analysis estimates that women experiencing early menopause (<45 years) have approximately 37% higher risk of all-cause dementia compared with women undergoing menopause at typical ages, although between-study heterogeneity was substantial.^14^ While previous studies demonstrate a link between timing of menopause in midlife and later life cognitive performance or dementia diagnosis, we lack an understanding of possible neurobiological links underlying this relationship.

Blood-based biomarkers of Alzheimer’s disease and related dementias (AD/ADRD) measured in representative epidemiologic cohorts offer an opportunity to evaluate *in vivo* dementia biology in a generalizable context.^15,16^ These emerging biomarkers, such as phosphorylated tau (ptau), glial fibrillary acidic protein (GFAP), neurofilament light chain (NfL), amyloid-beta isoforms, and growth differentiation factor-15 (GDF-15), correlate with AD-related neuropathology and are elevated in individuals with cognitive impairment and dementia.^17,18^ A growing literature links menopause-related endocrine biology to these biomarkers, with menopause stage and current levels of FSH and estradiol each associated with plasma and PET measures of tau, amyloid, and brain volume.^19–23^ However, this evidence comes largely from small, clinically-based, or single-site cohorts, leaving unresolved whether these biomarker-specific patterns replicate at the population level.

We aimed to provide the first nationally representative evidence on the association between age at spontaneous menopause and blood-based AD/ADRD biomarkers of pathophysiological processes. We examined a population-based cohort of postmenopausal women in the U.S., evaluating whether these associations differ by menopause type (spontaneous versus hysterectomy) and APOE ε4 carrier status.

## METHODS

### Participants

We used data from the U.S. Health and Retirement Study (HRS), a nationally representative longitudinal study of community-dwelling adults aged 50 years and older.^24^ The HRS, initiated in 1992, conducts biennial interviews that collect detailed information on health, socioeconomic conditions, and psychosocial factors. To maintain national representativeness, new birth cohorts are added approximately every six years.

In 2016, the HRS fielded a Venous Blood Study (VBS). All 2016 HRS respondents were invited to consent to an in-home blood draw, which 65% (n=9,934) completed. Among those who completed the VBS, a random subset of biospecimens was selected based on participants’ current or future eligibility for the Harmonized Cognitive Assessment Protocol. AD/ADRD biomarkers were assayed among this subset (n=4,539). Additional details have been published elsewhere.^17,25^

To ensure appropriate temporal ordering between exposure and outcome, we restricted our analytic sample to women who were at least 65 years of age at the VBS.

Given that HRS did not collect information regarding whether hysterectomy was performed with bilateral oophorectomy (“surgical menopause”) and that known changes in the hormone milieu occur even with hysterectomy alone^26^, we further excluded women who reported undergoing a hysterectomy before their age of menopause from the primary analytic sample. Secondary analyses evaluating associations between hysterectomy history and AD/ADRD biomarkers included 1,449 women. Consistent with prior studies,^22,27^ participants who reported menopause before age 20 or after age 60 were excluded from our analytic sample, as these ages are likely to reflect measurement error or atypical etiologies.

We further restricted the participants to those with complete data on biomarker outcomes, exposures, and covariates for a primary analytic sample of 893. Details on the analytic sample selection process are presented in **eFigure 1.** This study followed STROBE reporting guidelines for observational research.^28^

### Measures

#### Exposures

##### Primary

###### Menopausal timing

Menopausal status and age at spontaneous menopause were ascertained from HRS interview data collected between 2008 (when these questions were first administered) and 2016. Age at menopause was assessed biennially using the question: “About how old were you when you finished going through menopause, that is, had your last menstrual period?”

To minimize potential outcome-dependent misclassification, the first reported age of menopause was used in the primary analyses. Repeated reports of age at menopause demonstrated moderate-to-high within-person reliability (intraclass correlation coefficient [ICC] = 0.73).

##### Secondary

###### Hysterectomy status

At each wave, women self-reported a history of hysterectomy (yes/no) beginning in the 2008 HRS wave.

Bilateral oophorectomy and exogenous hormone use (e.g., contraceptives, hormone therapy) were not ascertained in the HRS.

#### Outcomes

Primary outcomes included neurofilament light chain (NfL), glial fibrillary acidic protein (GFAP), phosphorylated tau at threonine 181 (pTau-181), and growth differentiation factor 15 (GDF-15). Secondary outcomes included the ratio of plasma amyloid beta 42 to amyloid beta 40 (Aβ42/40) given its measurement in blood using immunoassays, which have demonstrated low concordance with gold standard measures of beta-amyloid biomarkers (e.g, CSF, PET) and modest disease-associated change.^29^

Blood samples were collected and processed using standardized protocols at the University of Minnesota Advanced Research and Diagnostic Laboratory. Details on sample collection, processing, and quality control have been described previously.^17,25^ Plasma NfL, GFAP, and Aβ42/40 were measured using the Neurology 4-plex Multiplex Simoa assay, and serum pTau-181 using a singleplex Simoa assay (Quanterix, Billerica, MA). Serum GDF-15 samples were processed on a Beckman Coulter Biomek NXp (Beckman Coulter, Fullerton, CA) and measured using a Simple Plex microfluidic immunoassay on the ELLA platform (ProteinSimple, San Jose, CA).

Given the right-skewed distribution of all biomarkers, outcomes were natural log-transformed. To facilitate interpretation and comparability across biomarkers, log-transformed values were additionally z-standardized. For interpretability, supplementary analyses report results using z-standardized values on the original (untransformed) scale. For all biomarkers except Aβ42/40, higher values reflect greater pathological burden.

#### Covariates

We included the following covariates that plausibly influence both age at menopause and AD/ADRD-related neurodegeneration ascertained at the same wave as the exposure (i.e., wave at which age at menopause was first reported): race and ethnicity (Non-Hispanic White, Non-Hispanic Black, Hispanic, Other racial/ethnic group), educational attainment (years), smoking status (never, past, current), live births (total number of children ever born), and marital status. We also adjusted for age at the 2016 assessment to capture cumulative aging up to the time of biomarker measurement.

In secondary models, we additionally adjusted for self-reported body mass index (kg/m^2^), hypertension, and diabetes, ascertained at the exposure wave. Sensitivity models additionally adjusted for kidney disease. These variables were considered separately from the primary adjustment set because they may lie on the causal pathway between menopause and neurodegeneration, and adjustment may cause unintended attenuation of the total effect estimates.^30^

Among a subsample (92%), APOE ε4 was genotyped using a TaqMan allelic discrimination SNP assay.^31^ Supplemental analyses evaluated effect modification by APOE ε4 carrier status (i.e., presence of at least one ε4 allele).

### Statistical Analysis

We first calculated descriptive statistics for the analytic sample. We then estimated associations between age at menopause and each biomarker outcome using generalized linear models, adjusted for the covariates described above. Age at menopause was modeled using restricted cubic splines with knots at 45, 50, and 55 years; clinical criteria and menopause timing distribution informed knot placement.^32^ We assessed evidence of nonlinearity using a Wald test of the nonlinear spline terms; as an alternative approach, we compared linear and spline models by likelihood ratio test and other fit statistics. When there was no evidence of nonlinearity, associations are presented as linear estimates. For outcomes with evidence of nonlinearity, we presented predicted marginal biomarker values across the observed range of age at menopause, along with the derivative (slope) estimates.

To assess the robustness of findings to differential inclusion in the ADRD biomarker sub-study, we conducted two sensitivity analyses. First, we applied truncated stabilized inverse probability of selection weights (IPWs) to account for differential participation in the ADRD biomarker study compared with the Venous Blood Study-eligible population. The second incorporated HRS survey weights to be nationally representative of the U.S. population represented by the 2016 HRS core study respondents born before 1960 (see **Appendix** for weighting details).

We conducted several additional sensitivity analyses. To reduce the influence of extreme values not reflective of a menopausal transition (e.g., primary ovarian insufficiency), we restricted to women reporting menopause at age ≥30 years. To account for the impact of kidney function on blood biomarker measurements, we additionally adjusted for estimated glomerular filtration rate (eGFR) at the time of the blood sample.^33^ To assess the robustness of nonlinear models, we fit restricted cubic splines with knots at the 10th, 50th, and 90th percentiles of the distribution, following recommended conventions for continuous variables with this sample size.^34^ We tested the average of all reported ages at menopause across biennial assessments. We additionally evaluated effect modification by APOE ε4 carrier status via multiplicative interaction terms and stratified models.

Secondary analyses evaluated the relationship between self-reported hysterectomy history and AD/ADRDs biomarkers. Analyses were conducted in Stata 18.

## RESULTS

### Descriptive characteristics

**Table 1** presents unweighted characteristics of the analytic sample. Among all women 65 years and older, 38.4% had a history of hysterectomy.

**Table 1.** Unweighted descriptive statistic [n (%) or mean (SD)], women who reported spontaneous menopause and participated in the U.S. Health and Retirement Study 2016 Venous Blood Neuropathology Study (n=893)

|  |  | Menopause Timing |  |  |
|---|---|---|---|---|
|  | **Overall** | **≤44** | **45-54** | **≥55** |
| N | 893 (100.0%) | 106 (11.9%) | 584 (65.4%) | 203 (22.7%) |
| Age (years) | 75.209 (7.208) | 75.604 (7.061) | 75.113 (7.360) | 75.281 (6.860) |
| Education (years) | 12.699 (3.326) | 11.340 (3.561) | 12.909 (3.296) | 12.803 (3.129) |
| Race and ethnicity |  |  |  |  |
| Non-Hispanic White | 641 (71.8%) | 64 (60.4%) | 433 (74.1%) | 144 (70.9%) |
| Non-Hispanic Black | 114 (12.8%) | 15 (14.2%) | 63 (10.8%) | 36 (17.7%) |
| Hispanic | 115 (12.9%) | 23 (21.7%) | 71 (12.2%) | 21 (10.3%) |
| Other | 23 (2.6%) | 4 (3.8%) | 17 (2.9%) | 2 (1.0%) |
| Smoking status |  |  |  |  |
| Never | 466 (52.2%) | 57 (53.8%) | 310 (53.1%) | 99 (48.8%) |
| Past | 332 (37.2%) | 32 (30.2%) | 218 (37.3%) | 82 (40.4%) |
| Current | 95 (10.6%) | 17 (16.0%) | 56 (9.6%) | 22 (10.8%) |
| Number of live births | 2.830 (1.922) | 3.094 (2.109) | 2.789 (1.939) | 2.808 (1.763) |
| Marital status |  |  |  |  |
| Married | 510 (57.1%) | 59 (55.7%) | 335 (57.4%) | 116 (57.1%) |
| Separated/divorced | 145 (16.2%) | 16 (15.1%) | 96 (16.4%) | 33 (16.3%) |
| Widowed | 211 (23.6%) | 27 (25.5%) | 135 (23.1%) | 49 (24.1%) |
| Never married | 27 (3.0%) | 4 (3.8%) | 18 (3.1%) | 5 (2.5%) |
| Self-reported BMI (kg/cm3) | 28.419 (6.165) | 27.470 (6.019) | 28.329 (5.862) | 29.186 (6.997) |
| Self-reported diabetes | 161 (18.0%) | 19 (17.9%) | 99 (17.0%) | 43 (21.2%) |
| Self-reported hypertension | 500 (56.0%) | 62 (58.5%) | 314 (53.8%) | 124 (61.1%) |
| APOE ε4 carrier status<sup>*a*</sup> | 199 (24.2%) | 18 (19.4%) | 133 (24.8%) | 48 (24.9%) |
| Age at menopause | 50.225 (5.223) | - | - | - |

*a*. APOE genotype is only available in a subsample (n=823) from which prevalence estimates are generated from.
Note: Mean (SD) for neurofilament light chain: 30.629 (SD ± 34.272); glial fibrillary acidic protein: 139.248 (SD ± 80.120); amyloid-beta 42/40 ratio: 0.066 (SD ± 0.016); phosphorylated tau 181: 2.247 (SD ± 2.520); growth differentiation factor: 1852.878 (SD ± 1344.118). With the exception of age, all covariates were ascertained at the same wave as the exposure (i.e., the wave at which age at menopause was first reported).
Abbreviations: SD, standard deviation.
Source: 2016 Health and Retirement Study, Venous Blood Study participants. Descriptive statistics are unweighted.

Among women without a history of hysterectomy (primary analytic sample), the mean age at blood collection was 75.2 years (standard deviation [SD]=7.2), mean educational attainment was 12.7 years (SD=3.3), and 72% identified as Non-Hispanic White. Mean age at menopause was 50.2 years (SD=5.2), and women were, on average, 25.0 years postmenopausal (SD=9.0). Characteristics were similar after applying IPWs to account for differential participation (**eTable 1**).

### Associations Between Menopausal Timing and AD/ADRD Biomarkers

Menopausal timing was significantly associated with NfL (Wald test: F[2,881] = 5.07, p = 0.007), and the association departed significantly from linearity (p = 0.002) (**Figure 1**). Covariate-adjusted predicted NfL values followed a nonlinear pattern, showing highest levels in women who reached menopause in their mid-to-late 40s and lower levels thereafter. Consistent with this pattern, average slopes were 0.010 SD units/year (95% CI: −0.007, 0.027) from menopausal ages 45–50 and −0.031 SD units/year (95% CI: −0.051, −0.011) from menopausal ages 50–55. Timing-specific estimates are shown in **eTable 2**.

**Figure 1.**
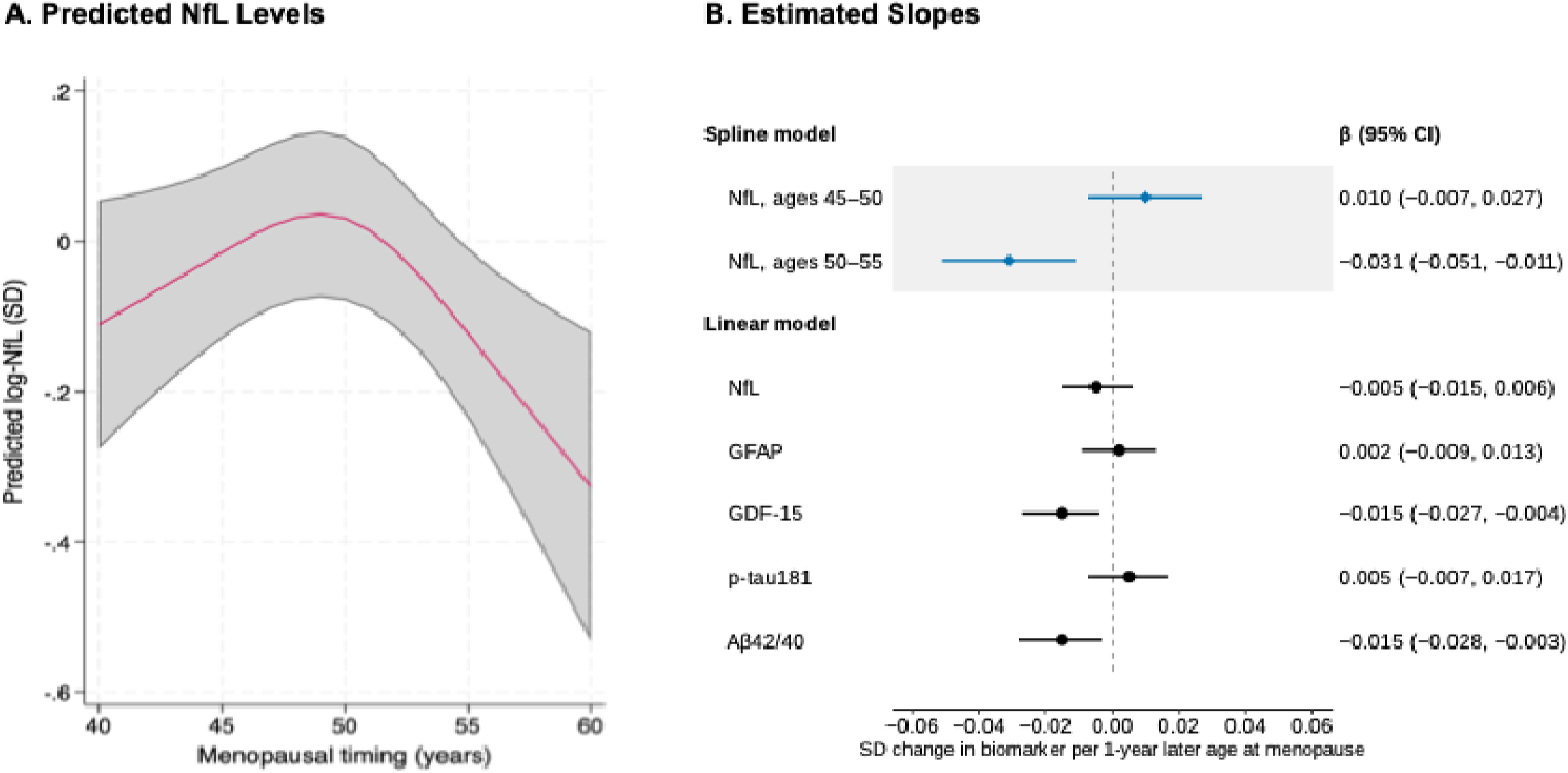
Association of menopausal timing with AD/ADRD-related biomarkers. A) Predicted NfL levels according to menopausal timing from the covariate-adjusted restricted cubic spline model, knot placement at ages 45,50, and 55. The shaded area indicates the 95% CI. B) Estimated associations of menopausal timing with AD/ADRD-related biomarkers. For NfL, estimates from the spline model represent the average change in predicted NfL per 1-year later age at menopause over the specified age intervals. For biomarkers modeled linearly, estimates represent the change in biomarker level per 1-year later age at menopause. Points indicate estimates and horizontal lines indicate 95% CIs. Biomarker values were standardized as log z-scores. Models were adjusted for age, education, race and ethnicity, smoking status, live births, and marital status. Abbreviations: Aβ42/40, amyloid-β 42 to 40 ratio; AD/ADRD, Alzheimer disease and Alzheimer disease–related dementias; GDF-15, growth differentiation factor 15; GFAP, glial fibrillary acidic protein; NfL, neurofilament light chain; p-tau181, phosphorylated tau 181. Source: Health and Retirement Study (2016; n=893)

There was no evidence of statistically significant nonlinearity in the association between menopausal timing and other AD/ADRDs biomarkers (**eFigure 2; eTable 3**). In linear models, later menopausal timing was associated with lower GDF-15 (*b* = −0.015, 95% CI: −0.027, −0.004) and lower Aβ42/40 (*b* = −0.015, 95% CI: −0.028, −0.003). Associations with GFAP (*b* = 0.002, 95% CI: −0.009, 0.013) and pTau-181 (*b* = 0.005, 95% CI: −0.007, 0.017) were not statistically significant (**Figure 1**).

### Sensitivity analyses

Results remained consistent after further adjustment for diabetes, hypertension, and obesity (**eTable 2**), as well as when utilizing z-score-standardized biomarkers without log transformation (**eTable 4**). APOE ε4 status did not significantly modify associations with biomarkers (**eFigures 3-4**). Stratified estimates suggested stronger associations between menopausal timing and AD/ADRD biomarkers in APOE ε4 carriers, most notably for pTau-181, though these differences did not reach statistical significance. Results were consistent after excluding ε2/ε4 carriers (**eTable 5**).

Models accounting for differential participation in the ADRD biomarker subsample produced consistent results, with significant nonlinear associations observed for NfL, and consistent overall associations observed for NfL, GDF-15, and Aβ42/40 (**Appendix 1; eTable 6**). Estimates weighted to be nationally representative followed similar directional patterns across all AD/ADRD-related biomarkers but were attenuated and less precise, particularly for Aβ42/40, for which associations were no longer statistically significant (**eTable 7**). Results were robust to the application of a more conservative threshold for allowable ages at menopause (**eTable 8**). Additional adjustment for kidney function^35^ resulted in consistent estimates (**eTable 9**). Results utilizing data-driven knot placement were consistent with those from the primary, clinically anchored specification (**eTable 10**), as were those utilizing the average value of the reported menopausal timing measure (**eTable 11**).

### Secondary analyses

Having a hysterectomy was associated with higher later life GDF-15 levels (*b* = 0.118, 95% CI: 0.015, 0.220); associations with other AD/ADRD biomarkers were not statistically significant (**eTable 12; Table 2**).

**Table 2.** Association between hysterectomy status and AD/ADRD-related biomarkers values (log z-score) (n=1,449)

| **Exposure** | NfL<br>β (95% CI) | GFAP<br>β (95% CI) | GDF-15<br>β (95% CI) | pTau-181<br>β (95% CI) | Aβ42/40<br>β (95% CI) |
|---|---|---|---|---|---|
| Hysterectomy (vs. no) | 0.021<br>(−0.072 to 0.113) | 0.027<br>(−0.069 to 0.124) | 0.118<br>(0.015 to 0.220) | 0.083<br>(−0.017 to 0.183) | 0.029<br>(−0.084 to 0.141) |

Abbreviations: Aβ42/40, amyloid-β42/40 ratio; GFAP, glial fibrillary acidic protein; GDF-15, growth differentiation factor 15; NfL, neurofilament light chain.
Notes: Estimates are β coefficients from multivariable linear regression models with log-transformed, z-standardized biomarker concentrations. Models were adjusted for age, race/ethnicity, education, smoking status, marital status, and number of live births.

## DISCUSSION

In this U.S. population-based study of women 65 years and older, we observed associations between menopausal timing and select blood biomarkers associated with dementia risk approximately 25 years following menopause. NfL, a marker of neuroaxonal injury not specific to AD pathology, was positively associated with menopausal timing at younger menopausal ages, before declining with progressively later menopausal timing. Later menopausal timing was otherwise linearly associated with lower GDF-15, a marker of immune aging, but not significantly associated with GFAP or pTau-181, markers of astrocyte reactivity and tau pathology. Collectively, these results provide population-level evidence of potential biological links underlying associations between midlife menopausal timing and later life neurodegenerative processes.

Several neuroendocrine mechanisms could underlie these associations, and/or menopausal timing itself may reflect a more central biological aging clock driving both reproductive and brain aging in tandem.^32^ Regarding the former, estradiol influences multiple neurobiological pathways relevant to dementia risk, including synaptic plasticity, neurotransmission, cerebrovascular regulation, mitochondrial metabolism, neurotrophic signaling, and neuroinflammation.^36–39^ Earlier menopausal timing may allow for expanded exposure or accelerated vascular, inflammatory, and neurodegenerative processes during midlife, when dementia-related biological changes may first emerge.^36,39,40^

The relationship between menopause timing and NfL reversed in direction at later ages. Interestingly, there was an unexpected, small positive association between menopausal timing and NfL at younger menopausal ages, which became progressively negative among women with later menopausal ages. The later-age pattern, in which more sustained exposure to youthful endogenous endocrine profiles (e.g., higher estradiol, low FSH) into later midlife was associated with lower NfL, is consistent with the data suggesting that prolonged estradiol exposure may confer some protection against neuroaxonal injury.^41^ Our findings, considered alongside prior observational evidence of a nonlinear relationship between age at menopause and dementia risk, suggest that associations may be complex and vary across the distribution of menopause age.^11^ Additional studies are needed to replicate our findings and further characterize the relationship between menopause and NfL.

Although associations with GFAP and pTau-181 showed patterns qualitatively similar to those observed for NfL, they were not statistically significant. This may reflect differences in biomarker specificity and/or limited statistical power, particularly for detecting nonlinear associations in spline-based models. Newer isoforms, particularly pTau-217, may be better suited to evaluating these associations because they appear less susceptible to regression-dilution bias.^42^ Replication in other nationally representative samples with improved measures is needed to determine whether these patterns reflect true but attenuated associations, as seen in prior literature.^43^

The association between later menopausal timing and a lower Aβ42/40 ratio, was unexpected and suggested that older age of menopause associated with amyloidogenic processing. This pattern contrasts with evidence suggesting that longer endogenous estrogen exposure is associated with lower amyloid-related pathology,^7,44,45^ and with cohort and PET-based studies linking earlier menopause to elevated Alzheimer’s risk and greater tau burden.^8,11^ ^3,46^ However, these results should be interpreted with caution given plasma Aβ42/40 estimates using immunoassays have been shown to suffer imprecision and the modest separation compared against levels of cerebral amyloid burden.^42^ Thus, these findings require replication in larger, diverse samples using amyloid assays with greater precision.

Later menopausal timing was linearly associated with lower GDF-15. GDF-15 is an emerging biomarker related to multiple organ aging, including the brain, and is elevated in vascular dementia, AD, and frontotemporal dementia.^47^ As a stress-responsive cytokine linked to mitochondrial dysfunction and systemic inflammation rather than AD-specific pathology, GDF-15 has been more strongly associated with vascular dementia than with Alzheimer’s disease in recent multi-cohort work.^47^ This aligns with a prior proteomic study, in which elevated GDF-15 was identified as the top protein correlate of earlier menopause.^23^ Considered alongside other reported findings, these results suggest that some of the biomarker differences associated with menopausal timing in this sample reflect systemic or vascular processes rather than AD-specific neurodegeneration, a distinction that future work with vascular imaging or cerebrovascular outcomes could help clarify.

We additionally found evidence from secondary analyses that a history of hysterectomy was associated with higher GDF-15 levels, but not with other biomarkers. We note that descriptively, hysterectomy history was very common among women 65+ in the HRS AD/ADRDs biomarker subsample, reflecting the high rate of hysterectomy among older birth cohorts in the U.S.^48,49^ Hysterectomy prevalence also varied by race and ethnicity, with the highest prevalence observed among non-Hispanic Black women, consistent with documented U.S. historical disparities in hysterectomy and sterilization practices.^50^ However, we interpret estimated associations with caution, as there is substantial heterogeneity in underlying hormonal trajectories among women with hysterectomy histories. While the HRS did not collect information on oophorectomy history, other population-based studies suggest that approximately half of women who underwent a hysterectomy did not also undergo bilateral oophorectomy.^51^

Several limitations merit consideration. First, menopausal timing was retrospectively self-reported. To minimize outcome-dependent misclassification, we used each participant’s earliest available report, which was less likely to be influenced by subsequent cognitive decline. Of note, we identified high reliability of self-reported menopause across visits (ICC = 0.7). Second, measurement error and random variability in blood-based biomarkers may have reduced statistical power to detect associations.^42,52^ However, results were consistent after additional adjustment for obesity and impaired kidney function, known sources of biomarker variability. Third, residual confounding remains possible, particularly from exogenous hormone use (i.e., oral contraceptives, hormone therapy), information not collected in the HRS.^53^ Additional relevant reproductive health factors (e.g., age of menarche, gynecologic conditions and surgeries outside of hysterectomy) were not captured in the present cohort and should be considered in future work. We note that the HRS only began asking participants about menopausal timing and hysterectomy in 2008, sixteen years after the first HRS wave was fielded. These measurement gaps reflect the broader landscape of research on women’s reproductive health histories and age-related outcomes.

In summary, this study provides novel, U.S. representative evidence that age at menopause associates with select blood-based biomarkers of AD/ADRD pathophysiology in a population-based sample of older U.S. women. Replication in other population-based studies, with richer data on reproductive health histories, including menopausal hormone therapy and surgical histories, including representation of diverse participants, will be essential for clarifying the clinical implications of these associations and identifying women who may benefit most from targeted prevention strategies. These findings underscore the importance of future work examining reproductive health to more deeply understand women’s later life dementia risk.

## Data Availability

All data produced in the present study are available upon reasonable request to the authors.

https://hrs.isr.umich.edu/data-products

## Notes

### Competing Interest Statement

The authors have declared no competing interest.

### Author Declarations

The University of Michigan Institutional Review Board gave ethical approval for this work (HUM00128220).

### Funding

- National Institute on Aging: T32AG049663, P01AG082653, RF1AG096165, K01AG075254
- Alzheimer's Association: AARG-25-1489464
- Wellcome Leap CARE program

