## Supplemental Material for "Age at Menopause and Dementia Risk: Blood-Based Biomarker Evidence from a US National Study"

**Supplement**

**eFigure 1**. Flowchart depicting participant selection into the primary analytical sample

**eTable 1**. Weighted descriptive statistics [n (%) or mean (SD)], women who reported spontaneous menopause and participated in the U.S. Health and Retirement Study 2016 Venous Blood Neuropathology Study

**eTable 2.** Estimated slopes for the association between menopausal timing and change in AD/ADRD-related biomarker values (log z-score) across menopausal timing

**eFigure 2.** Estimated slopes for the association between menopausal timing and change in AD/ADRD-related biomarkers (log z-score) among women who reported spontaneous menopause and participated in the U.S. Health and Retirement Study 2016 Venous Blood Neuropathology Study (n=893)

**eTable 3.** Comparing model fits between linear vs. non-linear specification of the AD/ADRD-related biomarkers (log z-score)

**eTable 4.** Estimated slopes for the association between menopausal timing and change in AD/ADRD-related biomarker values (z-score) across menopausal timing

**eFigure 3.** Association between menopausal timing and AD/ADRD-related biomarkers (log z-score) stratified by APOE status, linear functional form

**eFigure 4.** Estimated slopes for the association between menopausal timing and change in AD/ADRD-related biomarkers (log z-score) across menopausal timing stratified by APOE status among women who reported spontaneous menopause and participated in the U.S. Health and Retirement Study 2016 Venous Blood Neuropathology Study (n=893)

**eTable 5.** Effect modification by APOE ε4 status on the association between menopausal timing and AD/ADRD-related biomarkers (log z-score), excluding participants with the ε2/ε4 genotype (n=803)

**Appendix 1.** Construction and Interpretation of Inverse Probability Weights for AD/ADRD-related Biomarker Analyses

**eTable 6.** Weighted estimated slopes for the association between menopausal timing and AD/ADRD-related biomarkers (log z-score) across menopausal timing accounting for differential participation in the VBS ADRD Neuropathology Subsample

**eTable 7.** Population representative weighted estimated slopes for the association between menopausal timing and AD/ADRD-related biomarkers (log z-score) across menopausal timing

**eTable 8.** Estimated slopes for the association between menopausal timing and AD/ADRD-related biomarker values (log z-score) across menopausal timing among women with narrower menopausal ages (30-60)

**eTable 9.** Estimated slopes for the association between menopausal timing and AD/ADRD-related biomarker values (log z-score) across menopausal timing additionally adjusting for impaired kidney function

**eTable 10.** Estimated slopes for the association between menopausal timing and AD/ADRD-related biomarkers (log z-score) across menopausal timing with data-driven knot placement

**eTable 11.** Estimated slopes for the association between menopausal timing and AD/ADRD-related biomarkers (log z-score) across menopausal timing, utilizing average value of reported menopause age

**eTable 12**. Descriptive statistics [n (%) or mean (SD)] by hysterectomy status among women 65+ who participated in the U.S. Health and Retirement Study 2016 Venous Blood Neuropathology Study

Women who are 65+ who reported having spontaneous menopause

n= 962

Women (65+) with information on age at spontaneous menopause

n= 925

Women (65+) with all AD/ADRD biomarker information

n= 901

Final primary analytic sample

n= 893

Participated in the HRS Venous Blood Study (VBS), 2016
n= 9,924

Participated in the HRS VBS Neuropathology Study

n=4,539

Women who are 65+

n=1,542

Direct or proxy interviewees from the Health and Retirement Study (HRS), 2016

n= 20,192

10268 HRS interviewees who did not participate in the Venous Blood Study

5385 HRS-VBS participants did not participate in the neuropathology subsample

2997 participants who were younger than 65 years old and did not self-identify as women

580 women who did not have spontaneous menopause (e.g., surgical menopause, early hysterectomy)

37 participant missing exposure information

24 participants missing any outcome information

8 participants missing any covariate information

**eFigure 1**. Flowchart depicting participant selection into the primary analytical sample

| **eTable 1**. Weighted descriptive statistics [n (%) or mean (SD)], women who reported spontaneous menopause and participated in the U.S. Health and Retirement Study 2016 Venous Blood Neuropathology Study*^a,b^* | | | | |
| --- | --- | --- | --- | --- |
|  |  | **Unweighted** | **Weighted to account for differential participation** | **Weighted to the broader U.S. population of women 65+** |
| N |  | 893 (100.0%) | 889 (100.0%) | 15,791,973 (100.0%) |
| Age (years) |  | 75.209 (7.208) | 75.309 (7.252) | 74.303 (7.546) |
| Education (years) |  | 12.699 (3.326) | 12.649 (3.349) | 13.172 (3.133) |
| Race/ethnicity |  |  |  |  |
| Non-Hispanic White |  | 641 (71.8%) | 617 (69.4%) | 12,728,899 (80.6%) |
| Non-Hispanic Black |  | 114 (12.8%) | 131 (14.8%) | 1,233,327 (7.8%) |
| Hispanic |  | 115 (12.9%) | 118 (13.2%) | 1,424,524 (9.0%) |
| Other |  | 23 (2.6%) | 24 (2.6%) | 405,223 (2.6%) |
| Smoking status |  |  |  |  |
| Never |  | 466 (52.2%) | 464 (52.2%) | 8,137,380 (51.5%) |
| Past |  | 332 (37.2%) | 332 (37.3%) | 6,039,291 (38.2%) |
| Current |  | 95 (10.6%) | 93 (10.5%) | 1,615,302 (10.2%) |
| Number of live births |  | 2.830 (1.922) | 2.860 (1.942) | 2.612 (1.816) |
| Marital status |  |  |  |  |
| Married |  | 510 (57.1%) | 502 (56.4%) | 9,083,780 (57.5%) |
| Separated/divorced |  | 145 (16.2%) | 146 (16.4%) | 2,514,104 (15.9%) |
| Widowed |  | 211 (23.6%) | 214 (24.1%) | 3,401,989 (21.5%) |
| Never married |  | 27 (3.0%) | 27 (3.1%) | 792,100 (5.0%) |
| Self-reported BMI (kg/cm3) |  | 28.419 (6.165) | 28.468 (6.184) | 28.404 (6.223) |
| Self-reported diabetes |  | 161 (18.0%) | 163 (18.3%) | 2,689,718 (17.0%) |
| Self-reported hypertension |  | 500 (56.0%) | 503 (56.6%) | 8,385,026 (53.1%) |
| APOE ε4 carrier status *^c^* |  | 199 (24.2%) | 199 (24.3%) | 3,471,056 (23.7%) |
| Age at menopause |  | 50.225 (5.223) | 50.205 (5.262) | 50.345 (4.977) |
| *a*. Weights account for differential participation in the ADRD VBS study.  *b*. Descriptive statistics are weighted to be nationally representative  *c.* APOE genotype is only available in a subsample (n=823) from which prevalence estimates are generated from.  Abbreviations: SD, standard deviation.  Source: 2016 Health and Retirement Study, Venous Blood Study participants. Descriptive statistics are weighted. | | | | |

| **eTable 2.** Estimated slopes for the association between menopausal timing and change in AD/ADRD-related biomarker values (log z-score) across menopausal timing | | | | | | | |
| --- | --- | --- | --- | --- | --- | --- | --- |
|  | *Covariate set A* | | | | *Covariate set B* | | |
| Biomarker / Model | *Slope  (SD /year)* | LB  95% CI | Ub 95%CI | *Slope  (SD /year))* | | LB  95% CI | Ub 95%CI |
| NfL | | | | | | | |
| Linear | *-0.005* | -0.015 | 0.006 | *-0.006* | | -0.017 | 0.004 |
| Spline: Age 45 | *0.019* | 0.001 | 0.038 | *0.021* | | 0.003 | 0.039 |
| Spline: Age 50 | *-0.011* | -0.022 | 0.001 | *-0.014* | | -0.025 | -0.002 |
| Spline: Age 55 | *-0.041* | -0.066 | -0.015 | *-0.048* | | -0.074 | -0.023 |
| GFAP | | | | | | | |
| Linear | *0.002* | -0.009 | 0.013 | *0.002* | | -0.009 | 0.013 |
| GDF-15 | | | | | | | |
| Linear | *-0.015* | -0.027 | -0.004 | *-0.019* | | -0.030 | -0.008 |
| ptau-181 | | | | | | | |
| Linear | *0.005* | -0.007 | 0.017 | *0.003* | | -0.009 | 0.015 |
| Aβ42/40 ratio | | | | | | | |
| Linear | *-0.015* | -0.028 | -0.003 | *-0.014* | | -0.027 | -0.002 |
| Note. n = 893. Biomarker outcomes were log-transformed and z-scored. Linear models were fit for all biomarkers. For NfL, an additional model used restricted cubic splines with knots at ages 45, 50, and 55. Covariate Set A adjusted for age, race and ethnicity, education, number of live births, smoking status, and marital status. Covariate Set B additionally adjusted for body mass index, hypertension, and diabetes. β = SD change in biomarker per 1-year increase in age at menopause. Slope for spline models reflect dy/dx = instantaneous SD change in biomarker per 1-year increase in age at menopause, evaluated at the specified age. *P*_overall_ = P value from a joint Wald test of all spline terms, testing whether age at menopause is associated with the biomarker *P*_nonlinear_ = P value from a Wald test of the nonlinear spline term, testing whether the association departs from linearity. For NfL *P*_overall_ = 0.007 and *P*_nonlinea_= 0.002 (Covariate Set A); *P*_overall_ = 0.001 and *P*_nonlinea_ < 0.001 (Covariate Set B).  Abbreviations: NfL = Neurofilament light chain; GFAP = Glial fibrillary acidic protein; GDF-15 = Growth differentiation factor 15 ; p-tau181 = Phosphorylated tau 181; Aβ42/40 = Amyloid-beta 42/40 ratio. 95% confidence intervals shown in brackets. | | | | | | | |

| A) NfL   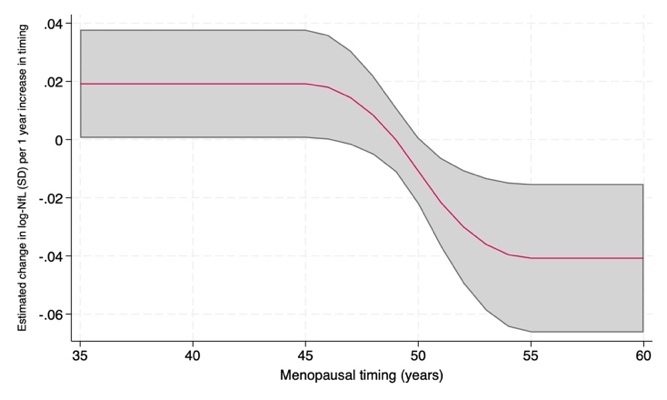  p = 0.002 for non-linearity  p = 0.01 for overall association | B) GFAP   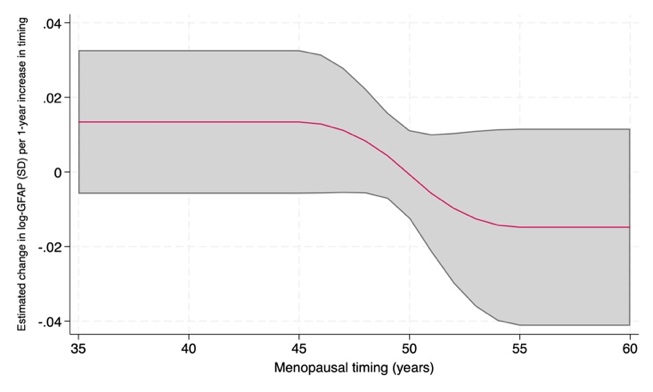  p = 0.16 for non-linearity p = 0.35 for overall association | C) GDF-15   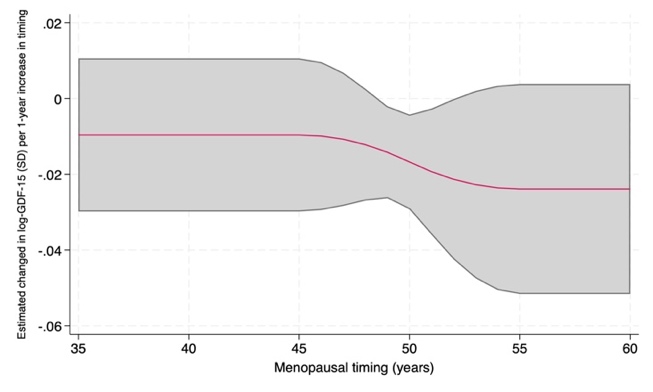  p = 0.50 for non-linearity  p = 0.03 for overall association |
| --- | --- | --- |
| D) ptau-181   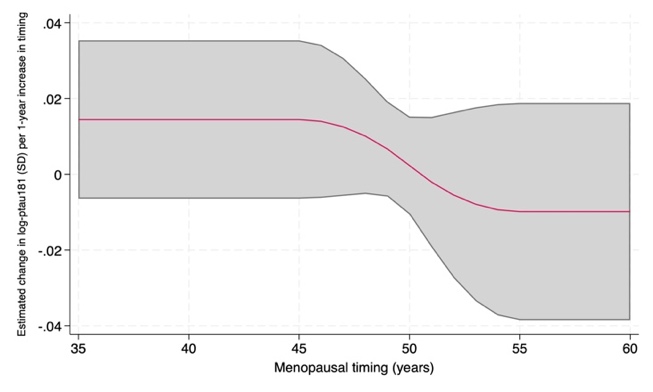  p = 0.27 for non-linearity p = 0.40 for overall association | E) Aβ42/40   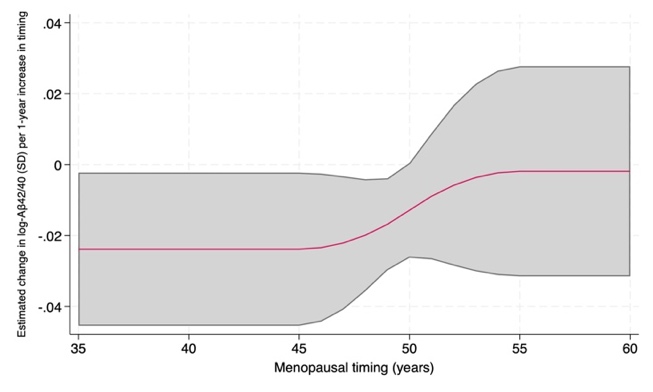  p = 0.33 for non-linearity p = 0.04 for overall association |  |
| **eFigure 2.** Estimated slopes for the association between menopausal timing and change in AD/ADRD-related biomarkers (log z-score) among women who reported spontaneous menopause and participated in the U.S. Health and Retirement Study 2016 Venous Blood Neuropathology Study (n=893). Note. The curve represents the estimated instantaneous rate of change in each biomarker (SD units) per 1-year increase in age at menopause, plotted continuously across ages; the shaded band represents the 95% confidence interval. Estimates derived from models adjusting for age, education, race and ethnicity, smoking status, live births, and marital status. Menopausal timing was modeled using restricted cubic splines with knots at ages 45, 50, and 55. *P*_overall_ is the p-value from a joint Wald test of all restricted cubic spline terms, assessing whether menopausal timing is associated with the biomarker. *P*_nonlinearity_ is the p-value from a Wald test of the nonlinear spline term(s), assessing whether the association departs from linearity. | | |

| **eTable 3.** Comparing model fits between linear vs. non-linear specification of the AD/ADRD-related biomarkers (log z-score) | | | | | | |
| --- | --- | --- | --- | --- | --- | --- |
| **Outcome** | **Model 1: Linear only** |  | **Model 2: Linear + Spline** |  |  | **LRT (M1 vs M2)** |
|  | β (95% CI), p | AIC / BIC | Overall p / Nonlinear (Wald) p | AIC / BIC |  | χ² (df), p |
| NfL | −0.005 (−0.015, 0.006), p=0.412 | 2257.7 / 2310.4 | **0.007** / **0.002** | 2250.1 / 2307.7 |  | 9.54 (1), p=**0.002** |
| GFAP | 0.002 (−0.009, 0.013), p=0.696 | 2312.3 / 2365.0 | 0.349 / 0.163 | 2312.3 / 2369.9 |  | 1.98 (1), p=0.160 |
| GDF-15 | −0.015 (−0.027, −0.004), p=**0.011** | 2396.1 / 2448.8 | **0.031** / 0.501 | 2397.6 / 2455.1 |  | 0.46 (1), p=0.498 |
| p-tau181 | 0.005 (−0.007, 0.017), p=0.434 | 2456.9 / 2509.6 | 0.398 / 0.267 | 2457.6 / 2515.2 |  | 1.25 (1), p=0.264 |
| Aβ42/40 | −0.015 (−0.028, −0.003), p=**0.018** | 2515.5 / 2568.3 | **0.038** / 0.332 | 2516.6 / 2574.1 |  | 0.95 (1), p=0.329 |
| Note: Models adjusted for age, race and ethnicity, education, live births, smoking status, and marital status. Menopausal age was modeled as a restricted cubic spline (knots at 45, 50, 55 years) in Model 2; Model 1 includes menopausal age linearly. Outcomes are log-transformed and z-scored (n=893); all models are linear regressions. "Overall p" tests the joint significance of the menopausal age terms; "Nonlinear (Wald) p" tests the nonlinear spline term(s) alone, indicating departure from linearity. Bold indicates p<0.05.  Abbreviations: AIC, Akaike information criterion; Aβ42/40, amyloid-β42/40 ratio; BIC, Bayesian information criterion; CI, confidence interval; df, degrees of freedom; GDF-15, growth differentiation factor 15; GFAP, glial fibrillary acidic protein; LRT, likelihood ratio test; NfL, neurofilament light chain. | | | | | | |

| **eTable 4.** Estimated slopes for the association between menopausal timing and change in AD/ADRD-related biomarker values (z-score) across menopausal timing | | | | |
| --- | --- | --- | --- | --- |
| Biomarker / Model | *Slope  (SD /year)* | LB  95% CI | Ub 95%CI | *p*_Non-linear_ |
| NfL | | | | |
| Linear | *-0.002* | -0.014 | 0.011 |  |
| Spline: Age 45 | *0.025* | 0.004 | 0.046 | 0.002 |
| Spline: Age 50 | *-0.009* | -0.022 | 0.004 |  |
| Spline: Age 55 | *-0.042* | -0.071 | -0.014 |  |
| GFAP | | | | |
| Linear | *0.001* | -0.010 | 0.013 |  |
| GDF-15 | | | | |
| Linear | *-0.015* | -0.027 | -0.003 |  |
| ptau-181 | | | | |
| Linear | *0.006* | -0.006 | 0.019 |  |
| Aβ42/40 ratio | | | | |
| Linear | *-0.015* | -0.028 | -0.003 |  |
| Note. n = 893. Biomarker outcomes were z-scored. Linear models were fit for all biomarkers. For NfL, an additional model used restricted cubic splines with knots at ages 45, 50, and 55. Covariate Set A adjusted for age, race and ethnicity, education, number of live births, smoking status, and marital status. β = SD change in biomarker per 1-year increase in age at menopause. Slope for spline models reflect dy/dx = instantaneous SD change in biomarker per 1-year increase in age at menopause, evaluated at the specified age. *P*_overall_ = P value from a joint Wald test of all spline terms, testing whether age at menopause is associated with the biomarker *P*_nonlinear_ = P value from a Wald test of the nonlinear spline term, testing whether the association departs from linearity. For NfL *P*_overall_ = 0.008 and *P*_nonlinea_= 0.002.  Abbreviations: NfL = Neurofilament light chain; GFAP = Glial fibrillary acidic protein; GDF-15 = Growth differentiation factor 15 ; p-tau181 = Phosphorylated tau 181; Aβ42/40 = Amyloid-beta 42/40 ratio. 95% confidence intervals shown in brackets. | | | | |

| A)  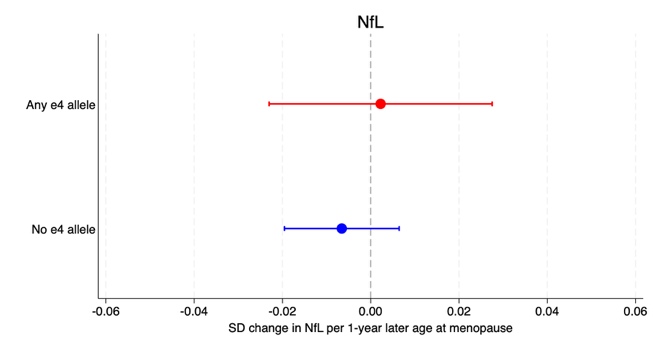  APOE ε4 × Age at menopause=  0.009 [95% CI: −0.020, 0.037]  p-interaction = 0.54 | B)  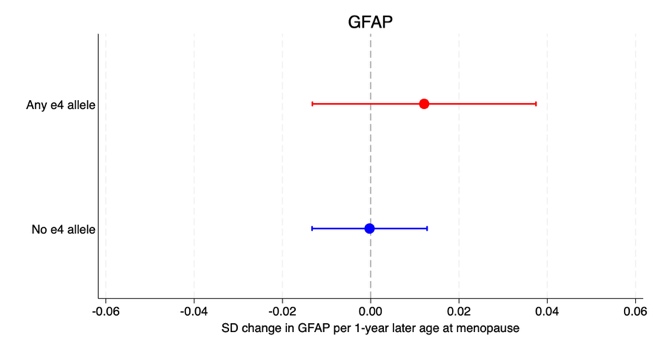  APOE ε4 × Age at menopause=  0.012 [95% CI: −0.016, 0.041]  p-interaction = 0.39 | C)  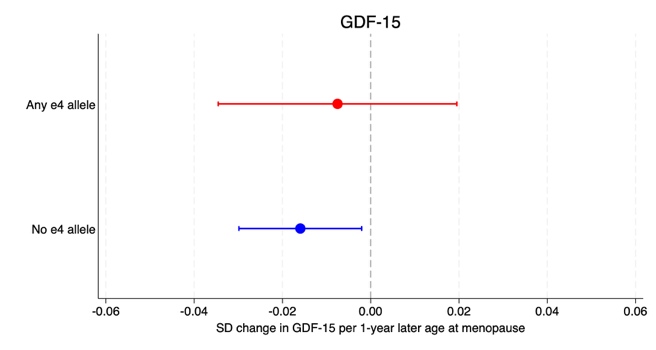APOE ε4 × Age at menopause=  0.008 [95% CI: −0.022, 0.039]  p-interaction = 0.59 |
| --- | --- | --- |
| D)  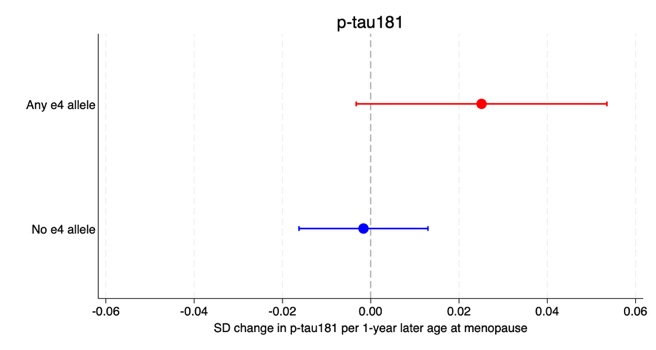  APOE ε4 × Age at menopause=  0.027 [95% CI: −0.005, 0.059]  p-interaction = 0.10 | E)  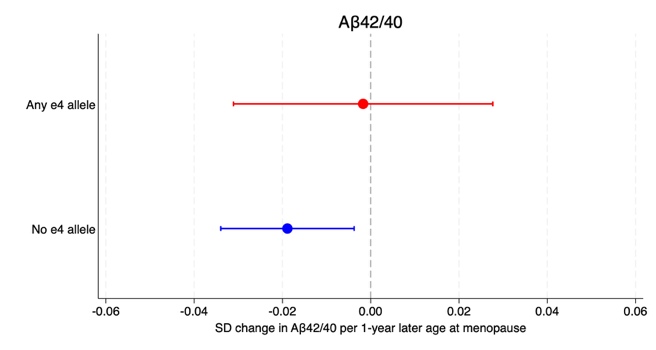  APOE ε4 × Age at menopause=  0.017 [95% CI: −0.016, 0.050]  p-interaction = 0.31 |  |
| **eFigure 3.** Association between menopausal timing and AD/ADRD-related biomarkers (log z-score) stratified by APOE status, linear functional form. Source: Health and Retirement Study (2016; n=823). Note. Marginal effects from multivariable linear regression models adjusting for age, education, race/ethnicity, smoking status, live births, and marital status. NfL = Neurofilament light chain; GFAP = Glial fibrillary acidic protein; Aβ42/40 = Amyloid-beta 42/40 ratio; p-tau181 = Phosphorylated tau 181; GDF-15 = Growth differentiation factor 15. APOE ε4 × Menopausal timing = interaction term between APOE ε4 carrier status and age at menopause. Bold indicates biomarkers for which a linear functional form was selected as the primary functional form. | | |

| A)  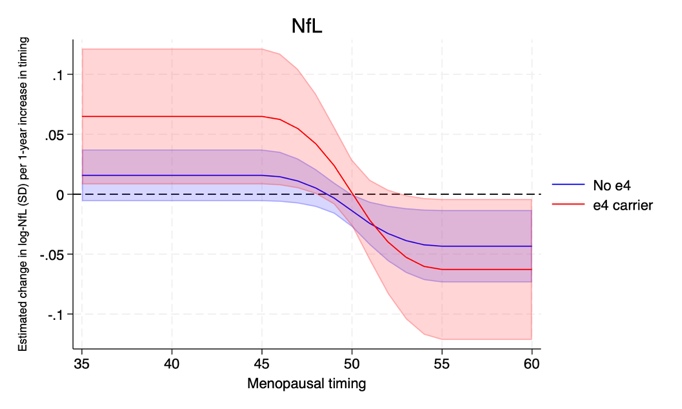  F[2,808] = 0.70; p-interaction = 0.50 | B)  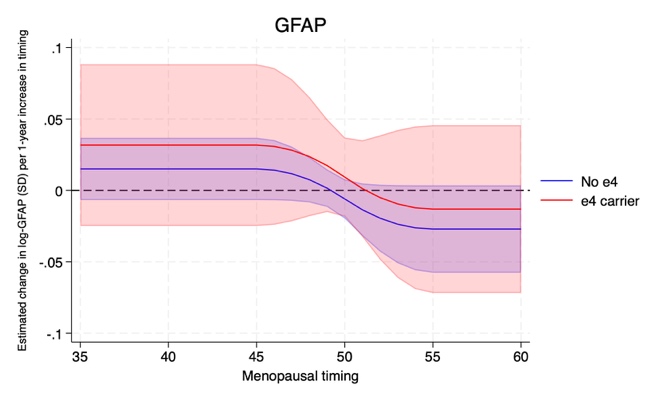  F[2,808] = 0.61; p-interaction = 0.54 | C)  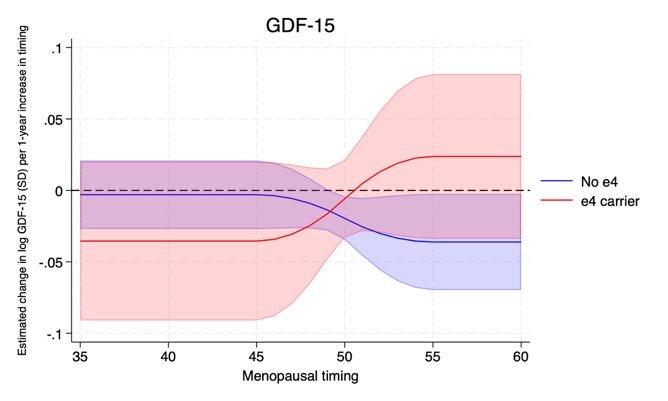  F[2,808] = 1.18; p-interaction = 0.31 |
| --- | --- | --- |
| D)  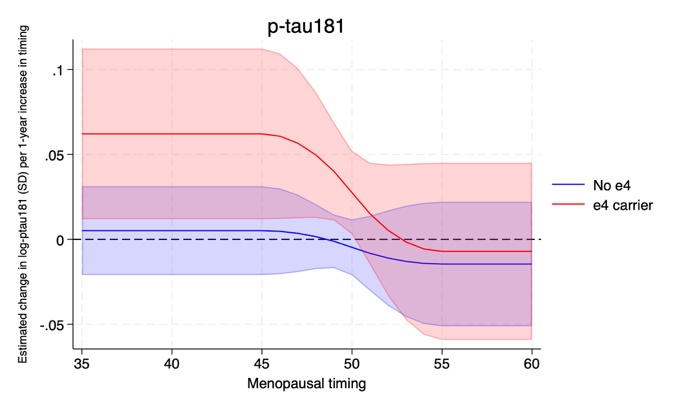  F[2,808] = 1.72; p-interaction = 0.18 | E)  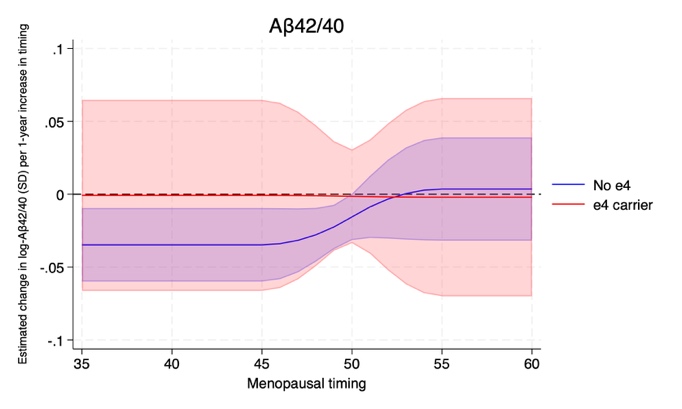  F[2,808] = 0.41; p-interaction = 0.66 |  |
| **eFigure 4.** Estimated slopes for the association between menopausal timing and change in AD/ADRD-related biomarkers (log z-score) across menopausal timing stratified by APOE status among women who reported spontaneous menopause and participated in the U.S. Health and Retirement Study 2016 Venous Blood Neuropathology Study (n=893). Note. Estimates derived from models adjusting for age, education, race/ethnicity, smoking status, live births, and marital status. Menopausal timing was modeled using restricted cubic splines with knots at ages 45, 50, and 55. Multiplicative interaction terms with restricted cubic splines were evaluated via joint Wald tests. Bold indicates biomarkers for which the restricted cubic spline was the primary functional form. | | |

| **eTable 5.** Effect modification by APOE ε4 status on the association between menopausal timing and AD/ADRD-related biomarkers (log z-score), excluding participants with the ε2/ε4 genotype (n=803) | | | | | | | | | | | | |
| --- | --- | --- | --- | --- | --- | --- | --- | --- | --- | --- | --- | --- |
| **Restricted cubic spline functional form** | | | | | | | | | | | | |
| 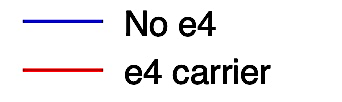  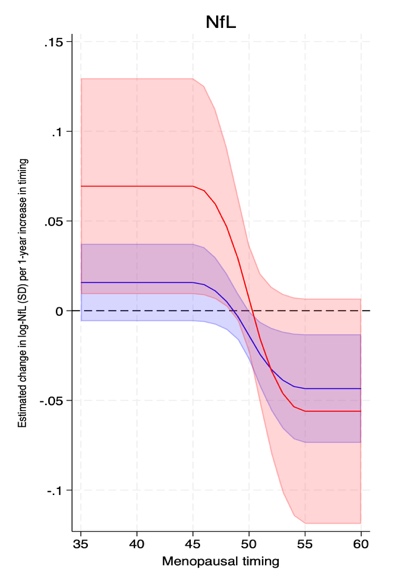  F[2,788] = 1.06;  p-interaction = 0.35 | | 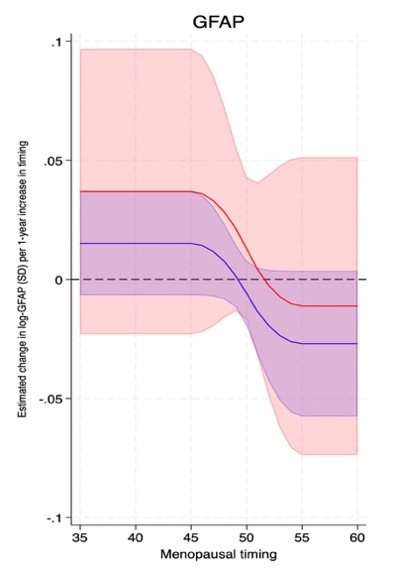  F[2,788] = 0.92;  p-interaction = 0.40 | | | 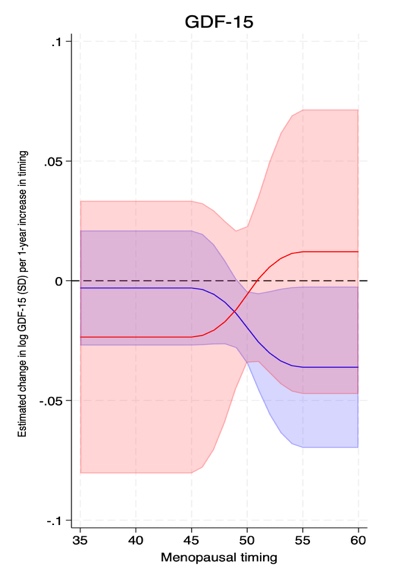  F[2,788] = 0.78;  p-interaction = 0.46 | | | 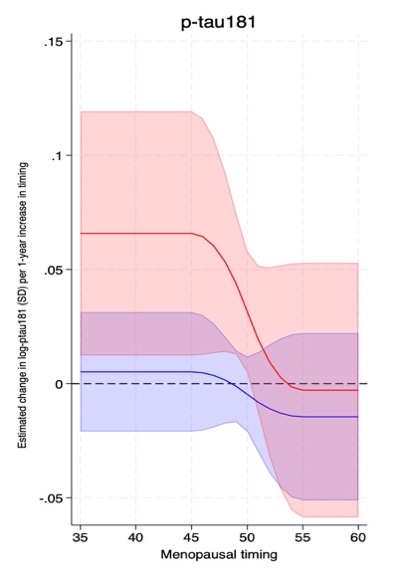  F[2,788] = 1.96;  p-interaction = 0.14 | | | 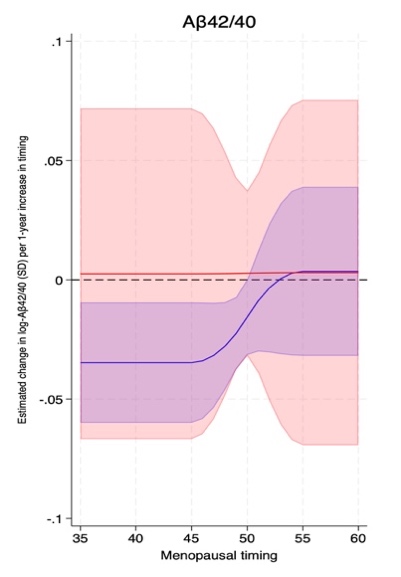  F[2,788] = 0.48;  p-interaction = 0.62 | |
| **Linear functional form** | | | | | | | | | | | | |
|  | NfL | | GFAP | | | GDF-15 | | | p-tau181 | | Aβ42/40 | |
|  | *β* | *95% CI* | *β* | *95% CI* | | *β* | *95% CI* | | *β* | *95% CI* | *β* | *95% CI* |
| APOE ε4 × Age at menopause | 0.014 | [−0.015, 0.043] | 0.017 | [−0.013, 0.046] | | 0.008 | [−0.024, 0.039] | | 0.031 | [−0.003, 0.064] | 0.019 | [−0.015, 0.053] |
| Note. NfL = Neurofilament light chain; GFAP = Glial fibrillary acidic protein; Aβ42/40 = Amyloid-beta 42/40 ratio; p-tau181 = Phosphorylated tau 181; GDF-15 = Growth differentiation factor 15. APOE ε4 × Menopausal timing = interaction term between APOE ε4 carrier status and age at menopause. Model adjusted for age, race/ethnicity, education, smoking, live births, and marital status. 95% confidence intervals shown in brackets. | | | | | | | | | | | | |

**Appendix 1.** Construction and Interpretation of Inverse Probability Weights for AD/ADRD-related Biomarker Analyses

**Overview**

Primary analyses were conducted without sampling weights. Because the analytic sample was restricted to women aged 65 years and older with complete AD/ADRD biomarker data, and our objective was to estimate associations within this analytic sample rather than produce population-representative estimates, sampling weights were not applied. Accordingly, unweighted regression coefficients directly estimate associations among the observed participants. To assess whether these associations were sensitive to differential participation in the U.S. Health and Retirement Study 2016 Venous Blood Neuropathology Study, and to additionally provide estimates that generalize to the broader population of women in the U.S., we conducted two weighted sensitivity analyses using inverse probability weights (IPWs). All weighted analyses used robust standard errors.

**Target Population**

The 2016 HRS ADRD Neuropathology sample is a subset of participants from the HRS 2016 Venous Blood Study (VBS) Sub-Sample. A central analytic decision in IPW construction is defining the target population; that is, the population to which weighted estimates are intended to generalize. We considered two target populations, corresponding to two distinct scientific questions:

- *Weight 1 – VBS-Eligible Women Aged 65 and Older.* This weight targets inference to women aged 65 and older, born prior to 1960, who were community-dwelling, non-proxy, and eligible for VBS participation in 2016 (i.e., excluding the Late Baby Boomer cohort, which entered the HRS sample in 2016 but was not eligible for VBS collection that wave). This is the population within which selection into the ADRD neuropathology sub-study operated. Restricting the target population to VBS-eligible women aged 65 and older, rather than extrapolating to the general U.S. population, allows this weight to correct specifically for differential participation in the neuropathology sub-study — matching our analytic sample exactly. We retain this alongside Weight 2 because the two weights address different questions: Weight 1 isolates the effect of sub-study selection, while Weight 2 (below) additionally incorporates the HRS complex sampling design to support inference to the general population. Comparing estimates across both provides a more complete picture of robustness than either alone.
- *Weight 2 – 2016 HRS Core Sample.* This weight targets inference to the broader population of women aged 65 and older, born prior to 1960, represented by the 2016 HRS core sample, constructed by scaling individual selection weights by the HRS 2016 respondent sampling weight (PWGTR), following the approach used to construct the official HRS VBS weights.

**Weight 1: Construction**

We constructed stabilized inverse probability of selection weights to account for differential participation in the ADRD neuropathology study relative to the VBS-eligible population of women aged 65 and older. The probability of ADRD neuropathology participation was estimated using logistic regression among VBS-eligible women aged 65 and older in the 2016 HRS (VBS16ELIG = 1) who met the eligibility criteria for VBS participation -- community-dwelling, born prior to 1960 (i.e., excluding the Late Baby Boomer cohort added to the HRS sample in 2016), non-proxy, and aged 65 or older (N = 5,539; ADRD neuropathology participants among this group, N= 1,494, 27%). Restricting the selection model to this subgroup directly (e.g., women only), rather than estimating it in a broader population and subsetting afterward, ensures the estimated selection propensities reflect the population within which our weighted estimates are intended to generalize. The model included the following covariates ${[ V}_{i}] :$age (continuous) and race/ethnicity as predictors of participation. Stabilized weights were constructed as the ratio of the marginal probability of ADRD participation to the predicted conditional probability of participation:

$\hat{W}_{i}=\hat{P}(ADRDVBS=1)/\hat{P}(ADRDVBS=1\mid V_{i} )$,

Weights were trimmed at the 99th percentile by Winsorizing values that exceeded this threshold, replacing them with the 99th percentile value. This approach limited the influence of extreme weights while retaining all observations in the analysis.

**Weight 2: Construction**

In the second sensitivity analysis, the HRS 2016 respondent sampling weight (PWGTR) was incorporated into the numerator of the IPW, following the guidance for the construction of the official HRS VBS Innovative Sub-Sample weights (VBSI16WGTRA/B). Specifically, weights were constructed as:

$\hat{W}_{i}=PWGTR_{i}/\hat{P}(ADRDVBS=1\mid V_{i})$,

As with Weight 1, weights were Winsorized at the 99th percentile, and post-stratified to the 2016 HRS target population of women aged 65 and older, born prior to 1960, by age and race/ethnicity. These weights target inference to the U.S. population of women aged 65 and older represented by the 2016 HRS core sample.

| **eTable 6.** Weighted estimated slopes for the association between menopausal timing and AD/ADRD-related biomarkers (log z-score) across menopausal timing accounting for differential participation in the VBS ADRD Neuropathology Subsample | | | | |
| --- | --- | --- | --- | --- |
| Biomarker / Model | *Slope  (SD /year)* | LB  95% CI | Ub 95%CI | *p*_Non-linear_ |
| NfL | | | | |
| Linear | *-0.004* | -0.015 | 0.006 |  |
| Spline: Age 45 | *0.020* | 0.002 | 0.037 | 0.002 |
| Spline: Age 50 | *-0.011* | -0.022 | 0.001 |  |
| Spline: Age 55 | *-0.041* | -0.067 | -0.015 |  |
| GFAP | | | | |
| Linear | *0.003* | -0.009 | 0.014 |  |
| GDF-15 | | | | |
| Linear | *-0.015* | -0.029 | -0.002 |  |
| ptau-181 | | | | |
| Linear | *0.005* | -0.006 | 0.016 |  |
| Aβ42/40 ratio | | | | |
| Linear | *-0.015* | -0.026 | -0.003 |  |
| Note. n = 893. Biomarker outcomes were log z-scored. Linear models were fit for all biomarkers. For NfL, an additional model used restricted cubic splines with knots at ages 45, 50, and 55. Weighted estimates account for differential participation in the VBS ADRD sub-study and include robust standard errors. Covariates included age, race and ethnicity, education, number of live births, smoking status, and marital status. β = SD change in biomarker per 1-year increase in age at menopause. Slope for spline models reflect dy/dx = instantaneous SD change in biomarker per 1-year increase in age at menopause, evaluated at the specified age. *P*_nonlinear_ = P value from a Wald test of the nonlinear spline term, testing whether the association departs from linearity.  Abbreviations: NfL = Neurofilament light chain; GFAP = Glial fibrillary acidic protein; GDF-15 = Growth differentiation factor 15 ; p-tau181 = Phosphorylated tau 181; Aβ42/40 = Amyloid-beta 42/40 ratio. 95% confidence intervals shown in brackets. | | | | |

| **eTable 7.** Population representative weighted estimated slopes for the association between menopausal timing and AD/ADRD-related biomarkers (log z-score) across menopausal timing | | | | |
| --- | --- | --- | --- | --- |
| Biomarker / Model | *Slope  (SD /year)* | LB  95% CI | Ub 95%CI | *p*_Non-linear_ |
| NfL | | | | |
| Linear | *-0.004* | -0.016 | 0.009 |  |
| Spline: Age 45 | *0.018* | -0.000 | 0.037 | 0.007 |
| Spline: Age 50 | *-0.009* | -0.022 | 0.004 |  |
| Spline: Age 55 | *-0.037* | -0.065 | -0.008 |  |
| GFAP | | | | |
| Linear | *0.003* | -0.010 | 0.016 |  |
| GDF-15 | | | | |
| Linear | *-0.015* | -0.028 | -0.003 |  |
| ptau-181 | | | | |
| Linear | *0.007* | -0.008 | 0.022 |  |
| Aβ42/40 ratio | | | | |
| Linear | *-0.009* | -0.023 | 0.006 |  |
| Note. n = 893. Biomarker outcomes were log z-scored. Linear models were fit for all biomarkers. For NfL, an additional model used restricted cubic splines with knots at ages 45, 50, and 55. Covariates included age, race and ethnicity, education, number of live births, smoking status, and marital status. β = SD change in biomarker per 1-year increase in age at menopause. Slope for spline models reflect dy/dx = instantaneous SD change in biomarker per 1-year increase in age at menopause, evaluated at the specified age. *P*_nonlinear_ = P value from a Wald test of the nonlinear spline term, testing whether the association departs from linearity.  Abbreviations: NfL = Neurofilament light chain; GFAP = Glial fibrillary acidic protein; GDF-15 = Growth differentiation factor 15 ; p-tau181 = Phosphorylated tau 181; Aβ42/40 = Amyloid-beta 42/40 ratio. 95% confidence intervals shown in brackets. | | | | |

| **eTable 8.** Estimated slopes for the association between menopausal timing and AD/ADRD-related biomarker values (log z-score) across menopausal timing among women with narrower menopausal ages (30-60) (n= 890) | | | | |
| --- | --- | --- | --- | --- |
| Biomarker / Model | *Slope  (SD /year)* | LB  95% CI | Ub 95%CI | *p*_Non-linear_ |
| NfL | | | | |
| Linear | *-0.007* | -0.018 | 0.005 |  |
| Spline: Age 45 | *0.019* | -0.002 | 0.041 | 0.004 |
| Spline: Age 50 | *-0.011* | -0.023 | 0.001 |  |
| Spline: Age 55 | *-0.041* | -0.067 | -0.015 |  |
| GFAP | | | | |
| Linear | *0.002* | -0.010 | 0.013 |  |
| GDF-15 | | | | |
| Linear | *-0.018* | -0.030 | -0.005 |  |
| ptau-181 | | | | |
| Linear | *0.003* | -0.010 | 0.016 |  |
| Aβ42/40 ratio | | | | |
| Linear | *-0.016* | -0.029 | -0.002 |  |
| Note. n = 890. Biomarker outcomes were log z-scored. Linear models were fit for all biomarkers. For NfL, an additional model used restricted cubic splines with knots at ages 45, 50, and 55. Covariates included age, race and ethnicity, education, number of live births, smoking status, and marital status. β = SD change in biomarker per 1-year increase in age at menopause. Slope for spline models reflect dy/dx = instantaneous SD change in biomarker per 1-year increase in age at menopause, evaluated at the specified age*. P*_nonlinear_ = P value from a Wald test of the nonlinear spline term, testing whether the association departs from linearity.  Abbreviations: NfL = Neurofilament light chain; GFAP = Glial fibrillary acidic protein; GDF-15 = Growth differentiation factor 15 ; p-tau181 = Phosphorylated tau 181; Aβ42/40 = Amyloid-beta 42/40 ratio. 95% confidence intervals shown in brackets. | | | | |

| **eTable 9.** Estimated slopes for the association between menopausal timing and AD/ADRD-related biomarker values (log z-score) across menopausal timing additionally adjusting for impaired kidney function | | | | |
| --- | --- | --- | --- | --- |
| Biomarker / Model | *Slope  (SD /year)* | LB  95% CI | Ub 95%CI | *p*_Non-linear_ |
| NfL | | | | |
| Linear | *-0.002* | -0.012 | 0.008 |  |
| Spline: Age 45 | *0.020* | 0.004 | 0.037 | 0.001 |
| Spline: Age 50 | *-0.008* | -0.018 | 0.002 |  |
| Spline: Age 55 | *-0.036* | -0.059 | -0.014 |  |
| GFAP | | | | |
| Linear | *0.003* | -0.008 | 0.014 |  |
| GDF-15 | | | | |
| Linear | *-0.013* | -0.024 | -0.002 |  |
| ptau-181 | | | | |
| Linear | *0.006* | -0.006 | 0.018 |  |
| Aβ42/40 ratio | | | | |
| Linear | *-0.015* | -0.028 | -0.003 |  |
| Note. n = 893. Biomarker outcomes were log z-scored. Linear models were fit for all biomarkers. For NfL, an additional model used restricted cubic splines with knots at ages 45, 50, and 55. Covariates included age, race and ethnicity, education, number of live births, smoking status, and marital status. β = SD change in biomarker per 1-year increase in age at menopause. Slope for spline models reflect dy/dx = instantaneous SD change in biomarker per 1-year increase in age at menopause, evaluated at the specified age. *P*_nonlinear_ = P value from a Wald test of the nonlinear spline term, testing whether the association departs from linearity.  Abbreviations: NfL = Neurofilament light chain; GFAP = Glial fibrillary acidic protein; GDF-15 = Growth differentiation factor 15 ; p-tau181 = Phosphorylated tau 181; Aβ42/40 = Amyloid-beta 42/40 ratio. 95% confidence intervals shown in brackets. | | | | |

| **eTable 10.** Estimated slopes for the association between menopausal timing and AD/ADRD-related biomarkers (log z-score) across menopausal timing with data-driven knot placement | | | | |
| --- | --- | --- | --- | --- |
| Biomarker / Model | *Slope  (SD /year)* | LB  95% CI | Ub 95%CI | *p*_Non-linear_ |
| NfL | | | | |
| Linear | *-0.005* | -0.015 | 0.006 |  |
| Spline: Age 45 | *0.019* | 0.002 | 0.037 | 0.002 |
| Spline: Age 50 | *-0.011* | -0.022 | 0.001 |  |
| Spline: Age 55 | *-0.041* | -0.067 | -0.015 |  |
| GFAP | | | | |
| Linear | *0.002* | -0.009 | 0.013 |  |
| GDF-15 | | | | |
| Linear | *-0.015* | -0.027 | -0.004 |  |
| ptau-181 | | | | |
| Linear | *0.005* | -0.007 | 0.017 |  |
| Aβ42/40 ratio | | | | |
| Linear | *-0.015* | -0.028 | -0.003 |  |
| Note. n = 893. Biomarker outcomes were log z-scored. Linear models were fit for all biomarkers. For NfL, an additional model used restricted cubic splines with knots at ages 45, 50, and 55. Covariates included age, race and ethnicity, education, number of live births, smoking status, and marital status. β = SD change in biomarker per 1-year increase in age at menopause. Slope for spline models reflect dy/dx = instantaneous SD change in biomarker per 1-year increase in age at menopause, evaluated at the specified age. *P*_nonlinear_ = P value from a Wald test of the nonlinear spline term, testing whether the association departs from linearity.  Abbreviations: NfL = Neurofilament light chain; GFAP = Glial fibrillary acidic protein; GDF-15 = Growth differentiation factor 15 ; p-tau181 = Phosphorylated tau 181; Aβ42/40 = Amyloid-beta 42/40 ratio. 95% confidence intervals shown in brackets. | | | | |

| **eTable 11.** Estimated slopes for the association between menopausal timing and AD/ADRD-related biomarkers (log z-score) across menopausal timing, utilizing average value of reported menopause age. | | | | |
| --- | --- | --- | --- | --- |
| Biomarker / Model | *Slope  (SD /year)* | LB  95% CI | Ub 95%CI | *p*_Non-linear_ |
| NfL | | | | |
| Linear | *-0.004* | -0.016 | 0.007 |  |
| Spline: Age 45 | *0.016* | -0.003 | 0.036 | 0.01 |
| Spline: Age 50 | *-0.010* | -0.022 | 0.003 |  |
| Spline: Age 55 | *-0.036* | -0.065 | -0.007 |  |
| GFAP | | | | |
| Linear | *0.002* | -0.010 | 0.014 |  |
| GDF-15 | | | | |
| Linear | *-0.020* | -0.033 | -0.008 |  |
| ptau-181 | | | | |
| Linear | *0.005* | -0.008 | 0.018 |  |
| Aβ42/40 ratio | | | | |
| Linear | *-0.014* | -0.027 | -0.001 |  |
| Note. n = 893. Biomarker outcomes were log z-scored. Linear models were fit for all biomarkers. For NfL, an additional model used restricted cubic splines with knots at ages 45, 50, and 55. Covariates included age, race and ethnicity, education, number of live births, smoking status, and marital status. β = SD change in biomarker per 1-year increase in age at menopause. Slope for spline models reflect dy/dx = instantaneous SD change in biomarker per 1-year increase in age at menopause, evaluated at the specified age. *P*_nonlinear_ = P value from a Wald test of the nonlinear spline term, testing whether the association departs from linearity.  Abbreviations: NfL = Neurofilament light chain; GFAP = Glial fibrillary acidic protein; GDF-15 = Growth differentiation factor 15 ; p-tau181 = Phosphorylated tau 181; Aβ42/40 = Amyloid-beta 42/40 ratio. 95% confidence intervals shown in brackets. | | | | |

| **eTable 12**. Descriptive statistics [n (%) or mean (SD)] by hysterectomy status among women 65+ who participated in the U.S. Health and Retirement Study 2016 Venous Blood Neuropathology Study*^a^* | | | |
| --- | --- | --- | --- |
|  |  | **Had spontaneous menopause (primary analytical sample)** | **Had a hysterectomy** |
| N |  | 893 (61.6%) | 556 (38.4%) |
| Age (years) |  | 75.209 (7.208) | 75.682 (7.288) |
| Education (years) |  | 12.699 (3.326) | 12.399 (2.746) |
| Race/ethnicity |  |  |  |
| Non-Hispanic White |  | 641 (71.8%) | 408 (73.4%) |
| Non-Hispanic Black |  | 114 (12.8%) | 96 (17.3%) |
| Hispanic |  | 115 (12.9%) | 52 (9.4%) |
| Other |  | 23 (2.6%) | 0 (0.0%) |
| Smoking status |  |  |  |
| Never |  | 466 (52.2%) | 281 (50.5%) |
| Past |  | 332 (37.2%) | 218 (39.2%) |
| Current |  | 95 (10.6%) | 57 (10.3%) |
| Number of live births |  | 2.830 (1.922) | 2.777 (1.642) |
| Marital status |  |  |  |
| Married |  | 510 (57.1%) | 249 (44.8%) |
| Separated/divorced |  | 145 (16.2%) | 88 (15.8%) |
| Widowed |  | 211 (23.6%) | 196 (35.3%) |
| Never married |  | 27 (3.0%) | 23 (4.1%) |
| Age at menopause |  | 50.225 (5.223) | n/a |
| *a.* Covariates assessed at the wave at which the exposure assessment was ascertained.  Abbreviations: SD, standard deviation.  Source: 2016 Health and Retirement Study, Venous Blood Study participants. Descriptive statistics are weighted. | | | |
